# Speed-emphasized arm training is consistent with improved feedforward control of rapid elbow flexion in chronic stroke

**DOI:** 10.64898/2026.10.02.26364609

**Authors:** Yannick Darmon, Gianni Hayrapetian, Shailesh Kantak, Victor R. Barradas, Gerald E. Loeb, Carolee Winstein, Emily R. Rosario, Nicolas Schweighofer

## Abstract

Feedforward control is impaired after stroke, but whether practice improves it in the chronic stage remains unclear. In fast single-joint movements, feedforward control scales initial acceleration to movement amplitude. Feedback-error-learning theory predicts that fast practice should promote recalibration. We tested whether speed-emphasized reach training improved this scaling in the more-affected arm. Thirty-six individuals with mild-to-moderate chronic stroke completed 2,080 reaches over four sessions emphasizing Speed (n = 17) or Accuracy (n = 19). At baseline, 3 days, and 1 month after training, participants performed single-joint elbow movements of three amplitudes without online visual feedback. The slope relating initial peak acceleration to target amplitude quantified feedforward scaling. At baseline, scaling was reduced on the more-affected side for extension (Δβ = −17.1 ± 3.5 s⁻², P < 0.001) and flexion (Δβ = −21.9 ± 3.2 s⁻², P < 0.001). Speed training increased flexion scaling at 3 days (Δβ = 14.7 ± 3.6 s⁻², P = 0.00029), with 14 of 17 participants improving. The group-by-session interaction showed a greater increase after Speed than Accuracy training (Δβ = 14.13 ± 4.9 s⁻², P = 0.0053). Scaling declined by 1 month (P = 0.02); no training-related changes occurred for extension. Peak acceleration occurred within 100 ms. Time to peak velocity, an index of pulse width, showed no group-specific training effect. Thus, speed-emphasized training produced a short-term improvement in early flexion acceleration scaling that transferred to an untrained single-joint task and was consistent with improved feedforward control. Transfer primarily reflected acceleration pulse-height rather than pulse-width modulation.

## Introduction

Approximately 65% of individuals post-stroke experience long-term limitations in upper-extremity function (Lum et al. 2009). Limitations in goal-directed reaching are particularly common and strongly correlate with impairment severity (Kamper et al. 2002; van Dokkum et al. 2014). Improvements in function and end-point kinematics may result from true recovery, defined as restoration of pre-stroke movement patterns; compensation, defined as the use of alternative movement strategies to accomplish a task; or a combination of both (Levin et al. 2009; Jones 2017; Nibras et al. 2021).

Rapid, accurate reaching depends on feedforward control to generate an initial motor command appropriate for the intended movement, because delayed sensory feedback cannot guide the earliest phase of movement. Supratentorial stroke can impair feedforward control, including the anticipatory specification of motor commands and the coordination of limb dynamics (Beer et al. 2000; Reinkensmeyer et al. 2003; Laczko et al. 2017; Raj et al. 2020). When the initial command is poorly specified, greater feedback-based correction is required to reach the target. Consistent with increased reliance on feedback control, reaching movements after stroke are typically slow and fragmented, with prolonged deceleration phases and multiple peaks in the velocity profile (Roby-Brami et al. 2003; McCrea and Eng 2005; Murphy et al. 2011; Stewart et al. 2014; Mottet et al. 2017). However, whether feedforward control can be improved through motor practice in the chronic stage after stroke remains unclear.

Feedback-error-learning theory provides a potential mechanism through which speed-focused practice could improve feedforward control. According to this theory, when an imperfect feedforward controller produces an inaccurate initial motor command, feedback controllers generate corrective motor commands that may also serve as teaching signals for calibrating the feedforward controller (Schweighofer, Arbib, et al. 1998; Kawato 1999; Kawato and Gomi 1992). Because faster movements place greater demands on feedforward control, a poorly calibrated controller should produce larger trajectory errors and, consequently, larger corrective feedback commands during fast than during slow movements. Repeated fast movements may therefore provide stronger teaching signals for recalibrating feedforward control.

The Phase 1 FAST randomized clinical trial tested this rationale by comparing speed- and accuracy-focused arm-movement training in individuals with chronic stroke (Darmon et al. 2025). Because the cerebellum is a key site for learning inverse models that support feedforward control, individuals with cerebellar stroke were excluded (Schweighofer, Spoelstra, et al. 1998; Kawato 1999). Across four sessions conducted over 1 week, participants completed 2,080 whole arm movements through complex tracks with the paretic arm. Speed and accuracy demands were manipulated using wide and narrow tracks, respectively, together with condition-specific performance feedback. At the 3-day post-test, speed training resulted in faster, smoother movements and more symmetric reach velocity profiles than accuracy training (Darmon et al. 2025). These changes were consistent with improved feedforward control but did not demonstrate it directly for three related reasons. First, although hand movements were planar and trunk motion was constrained, the task involved multiple arm degrees of freedom and therefore permitted compensatory inter-joint strategies. Second, joint torques generated during reaching were not estimated; estimating them from multidimensional arm kinematics would require a complex inverse-dynamics model that is sensitive to measurement noise and assumptions about model parameters and structure. Third, no simple quantitative relationship exists between end-point kinematics during multi-joint reaching and the underlying feedforward joint-torque commands. Thus, changes in smoothness and velocity-profile symmetry provided only indirect evidence of changes in feedforward control.

To address these limitations, the FAST trial also included a single-joint fast elbow movement task administered before and after training. Using a specially built apparatus that constrained movement to isolated elbow rotations, participants performed rapid horizontal flexion and extension of the elbow at different amplitudes about a low-viscosity, nearly frictionless axis. This task addressed each limitation of the reaching assessment. First, constraining the upper limb largely eliminated alternative inter-joint strategies, such that improvements more directly reflected changes in elbow control rather than compensation. Second, under these low-friction horizontal conditions, net elbow torque is approximately equal to the moment of inertia multiplied by angular acceleration. Thus, acceleration provides a biomechanically interpretable index of elbow torque without requiring a multidimensional inverse-dynamics model. Third, although both feedforward and feedback control contribute to the complete elbow movement, the initial acceleration peak occurs early in the movement, before delayed visual feedback can substantially influence it, although rapid proprioceptive contributions cannot be excluded. Because acceleration is directly related to torque in this task, the initial acceleration peak provides a relatively direct kinematic index of the initial feedforward torque command (Gordon and Ghez 1987; Kawato 1999).

For movements of different amplitudes, the initial motor command can be modulated in two complementary ways: by changing its magnitude (pulse-height control) or its duration (pulse-width control) (Gottlieb et al. 1990; Gordon and Ghez 1987). Pulse-height control is reflected in the scaling of initial peak acceleration with movement amplitude and has been associated with advance specification of movement extent, whereas pulse-width control is reflected in the duration of the initial positive acceleration pulse and may depend more strongly on sensory feedback (Sainburg and Schaefer 2004; Gordon and Ghez 1987). A well-calibrated feedforward controller should therefore scale the initial acceleration peak to the required movement amplitude. Reduced scaling indicates that the initial command is relatively insensitive to movement amplitude and may be compensated by prolonging the acceleration pulse or relying more on later feedback-based adjustments (Gordon and Ghez 1987; Sainburg and Schaefer 2004). Thus, the relationship between initial peak acceleration and movement amplitude provides an index of feedforward control, whereas time to peak velocity provides a complementary index of pulse width.

Using this fast elbow movement task, we tested the hypotheses that (1) before training, the more affected arm would show reduced scaling of initial peak acceleration with movement amplitude, indicating impaired feedforward control; and (2) speed training, compared with accuracy training, would increase this scaling, consistent with improved calibration of the feedforward controller.

## Materials and Methods

### Participants

Participants were recruited as part of the FAST clinical trial (Darmon et al. 2025), which included individuals with chronic stroke and mild-to-moderate upper extremity impairments. The present manuscript reports outcomes from an additional single-joint elbow movement assessment performed during the trial visits but not included in the primary FAST publication. Of the 42 participants enrolled in the trial, data from 36 participants (Speed group, n = 17; Accuracy group, n = 19) were available for the present analysis. Data from 6 participants were excluded because of an early protocol change that increased the number of target amplitudes and movements required to more accurately estimate target-dependent scaling (see below; n = 5) or equipment malfunctions during data collection (n = 1). Detailed descriptions of participant recruitment, inclusion and exclusion criteria, intervention procedures, and primary trial outcomes have been reported previously (Darmon et al. 2025). All participants provided written informed consent under a protocol approved by the Institutional Review Board of Casa Colina Hospital (IRB00002372).

Briefly, in the FAST trial, participants attended nine laboratory visits, including screening, evaluations of the less-affected and more-affected sides, four days of high-intensity upper-extremity training, a 3-day post-training evaluation, and a 1-month post-training evaluation. Participants were randomized to either a speed-biased or an accuracy-biased upper-extremity training intervention. Speed and accuracy demands were manipulated using wide and narrow tracks, respectively, together with condition-specific performance feedback. Both groups completed the same training dose, consisting of 2,080 complex reaching movements performed over four training sessions. During the four training sessions, participants in the Speed group completed the prescribed movements in less time, whereas participants in the Accuracy group accumulated a greater total movement distance. Participant characteristics and training metrics are summarized in Table 1. See (Darmon et al. 2025) for additional details.

**Table 1.** Summary of participant characteristics.

| Variable | Speed (n = 17) | Accuracy (n = 19) | P value |
| --- | --- | --- | --- |
| Age (years) | $57.5 \pm 3.6$ | $65.8 \pm 2.6$ | — |
| Female, n | 7 | 8 | — |
| UE-FM | $41.4 \pm 2.5$ | $42.8 \pm 1.7$ | — |
| Time since stroke (years) | $2.0 \pm 0.5$ | $3.1 \pm 1.1$ | — |
| Right hemiparesis, n | 9 | 9 | — |
| Training time (min) | $455 \pm 32$ | $715 \pm 29$ | *** |
| Training distance (m) | $845 \pm 35$ | $1,033 \pm 40$ | ** |

*Values are mean ± SE unless otherwise indicated.*
*P values are reported as significance levels: not significant —, $p < 0.05$ \*, $p < 0.01$ \*\*, $p < 0.001$ \*\*\*.*

### Fast Elbow Testing Task

During each assessment session, we measured elbow movement kinematics using a custom-built apparatus. The less-affected side was assessed once at baseline, whereas the more-affected side was assessed at baseline, 3-day post-training, and 1-month post-training. Participants were seated in a wheelchair with the trunk restrained by a four-point seatbelt. They were positioned as close as possible to the apparatus, which constrained movement to elbow flexion and extension (Figure 1A). The forearms were secured to the manipulandum with Velcro straps and maintained in a neutral pronation-supination position while the participant grasped the handle. The handles were adjusted so that the axis of rotation of the manipulandum was aligned with the elbow joint axis near the medial epicondyle. The forearm of the tested arm (less- or more-affected, depending on the session) was free to rotate about an axis under the elbow, whereas the other forearm was fixed. The height and width of the apparatus were adjusted so that the hand was positioned approximately 5 cm below the acromion and shoulder horizontal adduction was maintained at approximately 60°. Elbow angular displacement was measured using a potentiometer mounted at the axis of rotation. A monitor positioned in front of the participant displayed the virtual environment.

**Figure 1.**
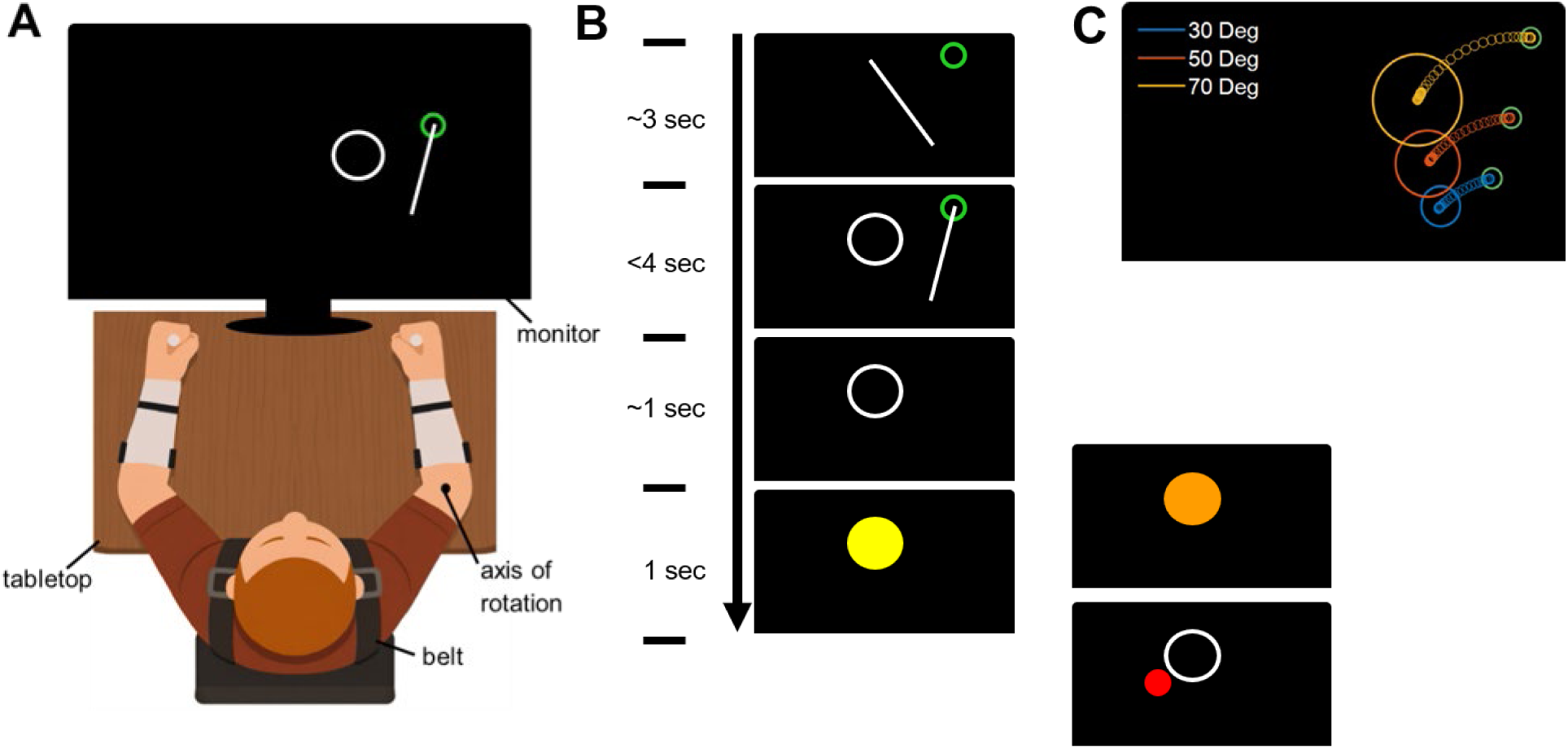
Experimental setup and task procedures. (A) Participant seated behind the apparatus, with trunk secured to the chair using a four-point seatbelt. The forearms were attached to the manipulandum via Velcro straps, and the virtual environment was displayed on a monitor positioned in front of the participant. Depending on the session (see text) one of the forearms was free to rotate around an axis of rotation under the elbow. (B) Experimental task sequence. Participant positioned the tip of the white line, representing the forearm, in the green circle representing the start position. Following a random wait time, the white circle representing the target appeared. Participant started reaching toward the target; once the white line left the start position, the white line and start position disappeared. Performance feedback was provided: a yellow disk indicated a successful reach within the time limit and target boundaries, an orange disk indicated the target was reached but exceeded the time limit, and a red dot indicated the reach ended outside the target. (C) Examples of hand trajectories for different target amplitudes. Target diameters increased with movement amplitude, with target diameters of 8.0 cm (70 deg, orange), 5.8 cm (50 deg, red), and 3.4 cm (30 deg, blue). For visualization, the example trajectories were shifted vertically to avoid overlap; during the experiment, all targets were positioned along the same arc defined by the virtual elbow position and forearm length.

Participants were instructed to perform fast and accurate elbow flexion or extension movements with amplitudes of 30°, 50°, or 70°. For flexion movements, the starting positions for the 30°, 50°, and 70° movements were 75°, 65°, and 55° of elbow flexion, respectively. The corresponding starting positions for extension movements were 115°, 125°, and 135° of elbow flexion (Figure 1C). A 10-cm white line representing the forearm was displayed on the screen and rotated as the participant moved the forearm (Figure 1B). The task required moving the line from a green circle representing the start position to a white circle representing the target (Figure 1B). After the participant maintained the white line within the start position for a randomized waiting period of 1 to 3 s, the target appeared. Once the forearm left the start position, the white line and start position disappeared (Figure 1B), so the movement was performed without online visual feedback. Target diameters of 3.4, 5.8, and 8.0 cm for the 30°, 50°, and 70° movements, respectively, maintained a constant index of difficulty (ID = 2), calculated according to Fitts’ law as ID = log₂(2D/W + 1), where D is movement amplitude and W is target diameter. Maintaining a constant index of difficulty minimized the influence of motor noise on movement planning (McCrea and Eng 2005), while increasing target width with movement amplitude permitted greater movement acceleration (Corcos et al. 1989).

In the second phase, participants performed the same movements while attempting to achieve a movement time below a predetermined threshold, corresponding to the 20th percentile of movement times recorded during the first phase for each movement direction and amplitude. These in-task computed movement times (as opposed to more accurately computed movement times for analyses; see below) were determined after each trial as the time required for the line tip to travel from the start position to the target. To limit the number of repeated trials, thresholds were bounded below at 0.10 s, 0.12 s, and 0.15 s for the small, medium, and large amplitudes, respectively. In addition to the feedback provided during the first phase, if the target was reached with a movement time exceeding the threshold, the target turned orange (Figure 1B), the message “Too slow” and the movement time were displayed, and the trial was repeated. Participants completed at least 10 flexion and 10 extension movements at each amplitude, for a minimum of 60 movements. Each unsuccessful trial could be repeated up to four times. Only successful trials from the second phase were included in the analyses, as peak acceleration scaling with movement amplitude is promoted by movement-speed requirements (Gottlieb et al. 1990). For 7 of 142 assessment sessions, participants were unable to complete the required number of successful trials within the time requirements; in these cases, we also included successful trials from the first phase. Two participants in the Accuracy group did not complete the 1-month assessment. The mixed-effects models used all available trials from completed assessments.

In addition, as part of the overall clinical trial, we assessed maximal voluntary torque during elbow flexion and extension at three different elbow positions (45°, 65°, and 85° of elbow flexion) (Johnson et al. 2026). At each position, participants performed two trials in either flexion or extension. During the first trial, participants generated a maximal flexion or extension torque against the apparatus. During the second trial, their best torque from the first trial was displayed as a target, and participants were instructed to move the cursor beyond this target to exceed their previous torque output.

### Data Analysis

Position signals from the potentiometer were digitized at 2 kHz using a USB analog-to-digital converter (USB-6259; National Instruments). Raw position signals were low-pass filtered at 8 Hz using a third-order, zero-phase, forward-backward Butterworth filter. The filtered position signal was differentiated to obtain velocity, and velocity was differentiated to obtain acceleration. Movement onset was defined as the first time the velocity exceeded 10% of the trial’s peak velocity, and movement end was defined as the first time the velocity fell below 10% of peak velocity after the peak. These thresholds were selected to avoid detecting small sub-movements while participants stabilized the forearm in the start position and were adapted from previously described threshold-based methods for detecting movement onset and end (Sainburg and Schaefer 2004). The initial acceleration peak was identified as the highest acceleration between movement onset and peak velocity. Time to the initial acceleration peak and time to peak velocity were calculated relative to movement onset. Because peak velocity marks the end of the initial positive acceleration pulse, time to peak velocity provided an index of pulse width (Sainburg and Schaefer 2004; Gottlieb et al. 1989). Movement time was calculated as the interval between movement onset and movement end.

### Statistical Analyses

To assess modulation of the initial acceleration peak as a function of target amplitude, we modeled peak acceleration at the trial level using linear mixed-effects models (R function *lmer* from the *lme4* package). For the more-affected side, peak acceleration was modeled as a function of target amplitude, assessment session (pretraining, 3 days post-training, and 1-month post-training), training group (Speed vs. Accuracy), and all interactions among these factors. Participant was included as a random effect, with a random intercept and random slopes for the target amplitude × session interaction to account for repeated measurements within individuals.

The primary outcome was the slope relating peak acceleration to target amplitude, which quantified the extent to which the initial acceleration peak scaled with movement amplitude. Changes in this scaling relationship across assessment sessions and training groups were assessed through interactions involving target amplitude. For this outcome, the pretraining-to-3-day difference between groups in slope change was the principal training-effect contrast; contrasts involving the 1-month assessment characterized retention.

We additionally analyzed peak acceleration magnitude, time to the initial acceleration peak, time to peak velocity, and movement time using linear mixed-effects models fit separately for flexion and extension movements, with assessment session, training group, and their interaction as fixed effects. Participant was included as a random effect, with random intercepts and random slopes for assessment session to account for repeated measurements within individuals. Time to the initial acceleration peak was used to assess whether the peak occurred within the early feedforward phase. Time to peak velocity and movement time were exploratory outcomes used to distinguish pulse-height modulation from changes in pulse width or overall movement duration.

Finally, to compare the less-affected and more-affected sides at baseline, data from the less-affected-side assessment and the pretraining more-affected-side assessment were analyzed together using the same linear mixed-effects model structures described above, with assessment session replaced by side (less-affected vs. more-affected). For the peak-acceleration scaling model, the random-effects structure included participant-specific random intercepts and random slopes for the target amplitude × side interaction. For the other kinematic outcomes, participant was included as a random effect, with random intercepts and random slopes for side. Differences between sides were assessed through the corresponding side effects and interactions.

Finally, maximal voluntary torque during flexion and extension was compared at baseline using a linear mixed-effects model with movement direction as a fixed effect and participant as a random effect.

Models were initially fitted using all available trials. Influential observations were identified using Cook’s distance; trials with unusually high peak acceleration values that exerted disproportionate influence on model estimates were excluded before refitting the models. For the baseline comparison between the less- and more-affected sides, 7 of 2,068 extension trials (0.34%) and 10 of 2,082 flexion trials (0.48%) were excluded. For the longitudinal analyses, 28 of 3,072 extension trials (0.91%) and 63 of 3,081 flexion trials (2.04%) were excluded. All reported estimates and statistical tests were obtained from the refitted models.

## Results

### Participant characteristics and training overview

The Speed (n = 17) and Accuracy (n = 19) groups did not differ in age, sex distribution, more-affected side, time since stroke, or upper-extremity impairment (all p > 0.05; Table 1). As expected, the training interventions produced distinct whole arm reaching behaviors. Participants in the Speed group completed the prescribed 2,080 movements across the four training sessions in significantly less time than those in the Accuracy group. The Speed group also generated significantly shorter hand-path distances during training, reflecting the straighter reaching trajectories associated with the speed-biased training paradigm (Table 1). See Darmon et al. (2025) for additional details about the effects of the training interventions on whole arm movement kinematics. Baseline strength also differed between movement directions, with lower maximal elbow-extension torque than maximal elbow-flexion torque (10.1 ± 1.0 vs. 12.3 ± 1.0 N·m; p < 0.0001).

### Initial acceleration peak timing

Elbow flexion and extension movements (Figure 1C) were characterized by a large initial acceleration peak followed by one or more smaller peaks (Figure 2A, B). For both movement directions and sides, the mean time to the initial acceleration peak was less than 100 ms, consistent with its use as an index of the initial feedforward motor command. For flexion, time to the initial acceleration peak was significantly shorter on the less-affected side (0.060 ± 0.004 s) than on the more-affected side (0.079 ± 0.005 s; Δ = −0.019 ± 0.004 s, p < 0.0001) at baseline. In the Speed group, time to the initial acceleration peak did not change from pretraining to the 3-day assessment, pretraining to the 1-month assessment, or the 3-day to the 1-month assessment (all p ≥ 0.31). Similarly, no significant changes were observed in the Accuracy group (all p ≥ 0.14), and no group by session interactions were detected (all p ≥ 0.36). For extension, time to the initial acceleration peak was also significantly shorter on the less-affected side (0.056 ± 0.004 s) than on the more-affected side (0.068 ± 0.005 s; Δ = −0.012 ± 0.005 s, p = 0.023) at baseline. No significant within-group changes were observed across sessions (p ≥ 0.15), and no group × session interactions were significant (p > 0.05).

**Figure 2.**
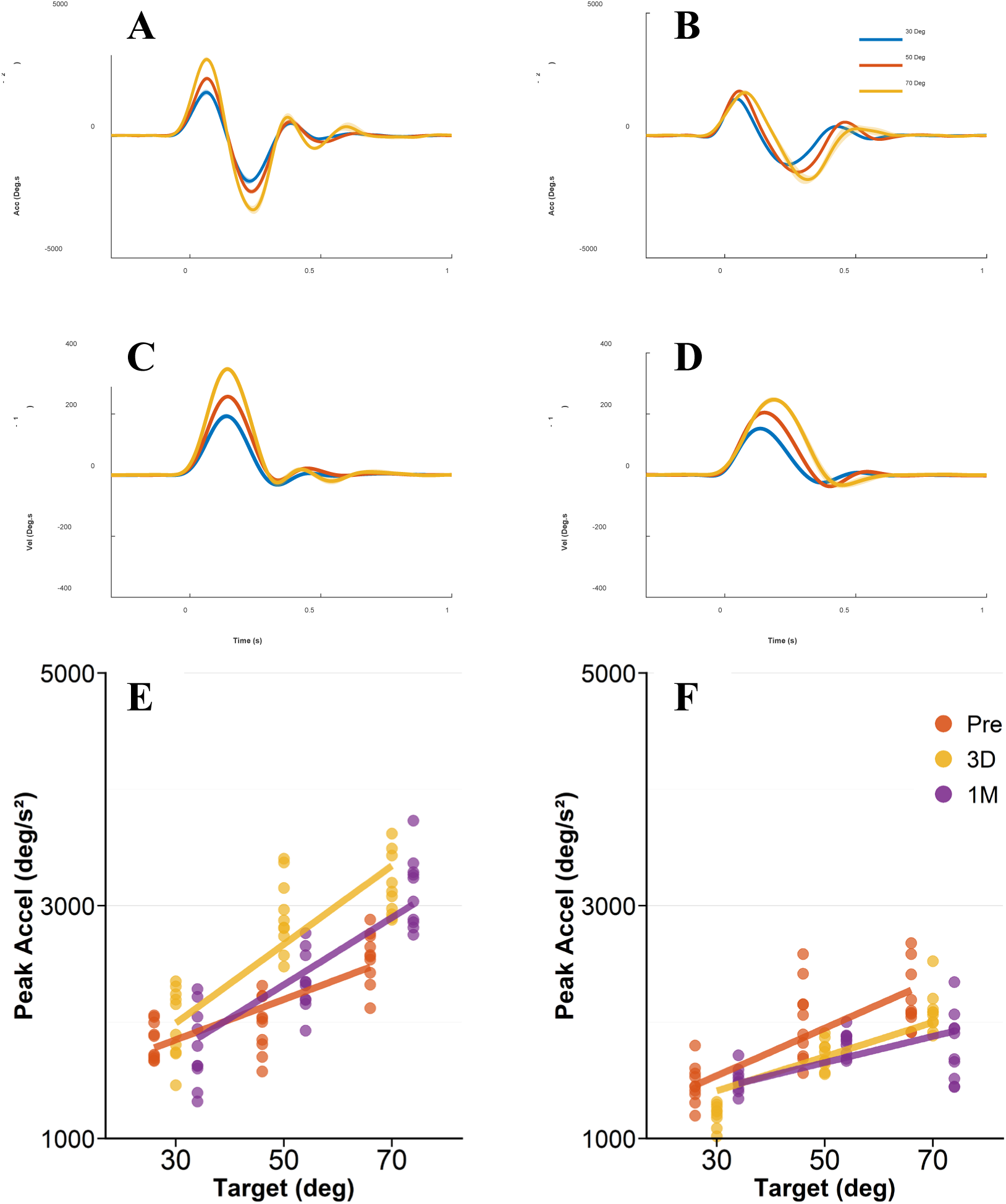
Representative acceleration and velocity profiles and peak acceleration scaling for flexion movements. (A–D) Mean ± SE elbow acceleration (A, B) and velocity (C, D) profiles from a participant exhibiting peak acceleration scaling with target amplitude (left panels; FT008, UE-FM = 48) and a participant exhibiting relatively constant peak acceleration across target amplitudes (right panels; FT022, UE-FM = 40) at the 1-month assessment. The yellow, red, and blue traces represent the average of 10 reaching movements toward targets requiring 70, 50, and 30 deg of elbow excursion, respectively. For display purposes, acceleration and velocity profiles were aligned to movement onset. (E, F) Representative examples of peak acceleration scaling for the more affected arm before (Pre; red), 3 days after (3D; yellow), and 1 month after (1M; purple) the intervention. Each dot represents the peak acceleration of a successful reaching movement at a given target amplitude. Lines represent the fitted relationships estimated from the three-way mixed-effects model. (E) Representative participant from the Speed group. (F) Representative participant from the Accuracy group.

### Peak acceleration scaling with target amplitude

The initial acceleration peak increased with target amplitude in some participants but changed little in others, yielding steeper and shallower individual scaling slopes, respectively. Figure 2E, F illustrates examples of scaling and little or no scaling across target amplitudes for the more-affected arm of two participants at the 1-month assessment.

#### Scaling was lower on the more-affected than on the less-affected side at baseline

On the less-affected side, peak acceleration scaled significantly with target amplitude for both extension (β = 37.8 ± 3.3 s⁻², p < 0.001) and flexion (β = 38.2 ± 2.8 s⁻², p < 0.001) at baseline. Scaling was significantly lower on the more-affected than on the less-affected side for both extension (Δβ = −17.1 ± 3.5 s⁻², p < 0.001) and flexion (Δβ = −21.9 ± 3.2 s⁻², p < 0.001). The reduction was numerically greater for flexion than extension, but the side-by-direction interaction was not significant (p = 0.18).

#### Speed, but not accuracy, training improved acceleration scaling for flexion movements

For flexion movements of the more-affected arm, peak acceleration scaled with target amplitude at baseline (β = 16.4 ± 2.1 s⁻², p < 0.001). Baseline scaling was numerically lower in the Speed (β = 13.7 ± 3.0 s⁻²) than Accuracy group (β = 20.9 ± 2.8 s⁻²), although the difference was not statistically significant (p = 0.09; Figure 3A). Speed training significantly increased the scaling slope from pretraining to the 3-day assessment (Δβ = 14.7 ± 3.5 s⁻², p = 0.00029; Figure 3A). In contrast, scaling did not change in the Accuracy group from pretraining to the 3-day assessment (p = 0.98). Accordingly, a significant group × session interaction was observed from pretraining to the 3-day assessment (Δβ = 14.13 ± 4.9 s⁻², p = 0.0053), indicating a greater short-term increase in scaling in the Speed group than in the Accuracy group (Figure 3B). This increase was observed in all but three participants (14 of 17) in the Speed group (Figure 3C), whereas only half (10 of 19) showed an increase in the Accuracy group (Figure 3C). In the Speed group, scaling subsequently decreased from the 3-day to the 1-month assessment (Δβ = −7.6 ± 3.3 s⁻², p = 0.022), indicating that some of the initial gains were not maintained at the 1-month follow-up (Figure 3A). The pretraining-to-1-month group × session contrast was Δβ = 9.2 ± 4.5 s⁻² (p = 0.048); two Accuracy participants lacked 1-month data for this contrast.

**Figure 3.**
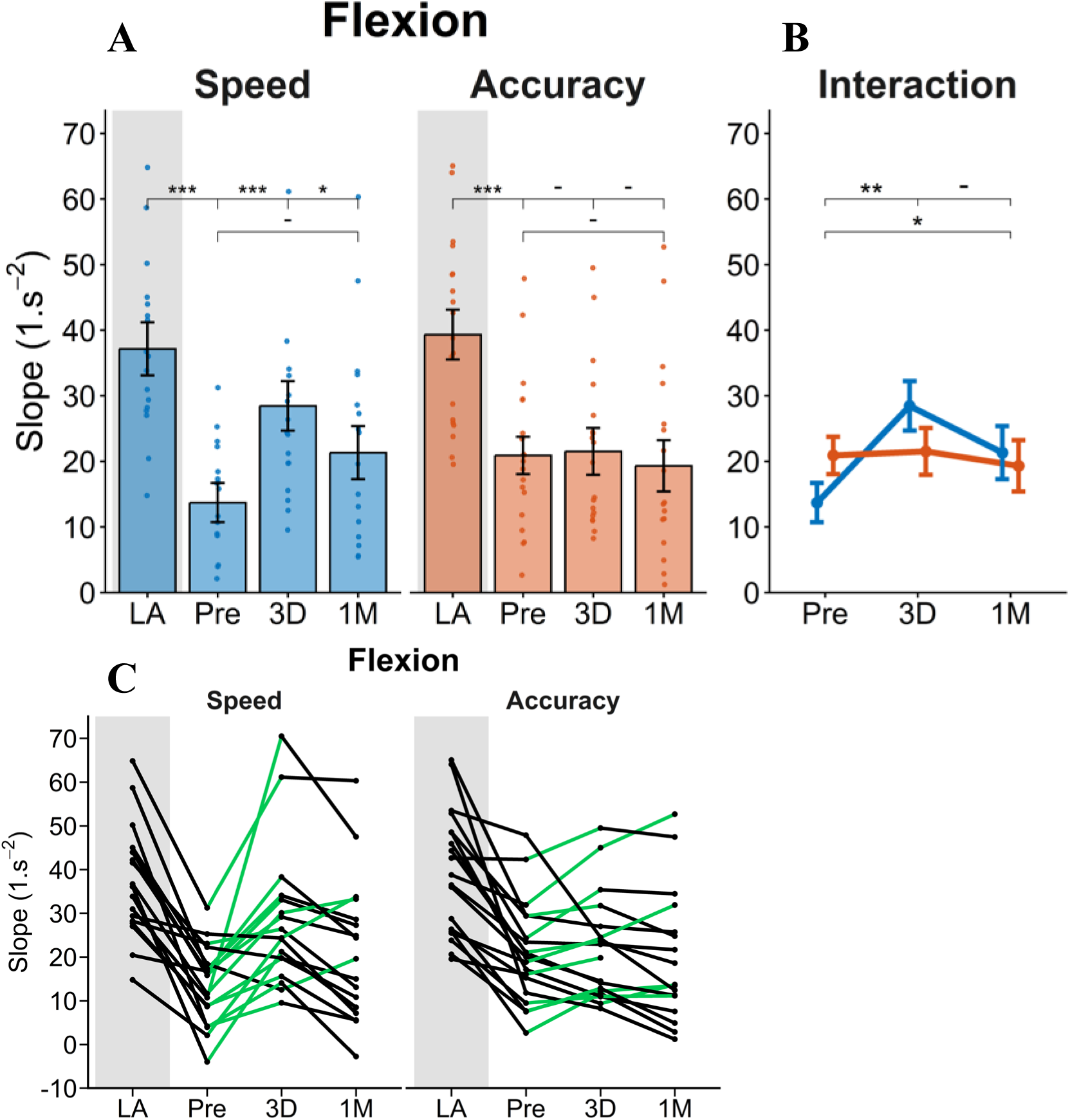
Individual and group changes in peak acceleration scaling slopes for the less affected side (LA) and more affected side: before training (Pre), 3 days after training (3D), and 1 month after training (1M). (A) Changes in scaling slopes across assessment sessions for the Speed and Accuracy groups. Each dot represents the fitted slope for an individual participant. Bars represent estimated marginal means ± SE. Horizontal bars indicate pairwise comparisons between assessment sessions. (B) Interaction effects between training group and assessment session for scaling slopes across each time interval (Pre to 3D, 3D to 1M, and Pre to 1M). Dots represent estimated marginal means ± SE, and horizontal bars indicate significance; P values are reported as significance levels: not significant -, p < 0.05 *, p < 0.01 **, p < 0.001 ***. (C) Individual participant changes in scaling slopes across assessment sessions. The shaded gray background represents the less affected side, whereas the white background represents the more affected side. Each dot represents the fitted scaling slope for an individual participant, and lines connect slope estimates from the same participant across assessment sessions. Green lines indicate an increase in slope, whereas black lines indicate a decrease in slope.

#### Flexion scaling changes primarily reflected pulse-height modulation

In an exploratory analysis, we examined time to peak velocity, which marks the end of the initial positive acceleration pulse and therefore indexes pulse duration, to determine whether the training-related increase in acceleration scaling was accompanied by pulse-width modulation. At baseline, time to peak velocity was significantly shorter on the less-affected side (0.143 ± 0.008 s) than on the more-affected side (0.169 ± 0.009 s; Δ = −0.026 ± 0.008 s, *p* = 0.002). Thus, at baseline, the more-affected arm combined reduced pulse-height scaling with a longer initial acceleration pulse.

In the Speed group, mean time to peak velocity decreased transiently from pretraining to the 3-day assessment, although not significantly (p = 0.07), and returned toward baseline by 1 month (p = 0.8). No significant changes were observed in the Accuracy group, and no group-by-session interactions were detected. Total movement time did not differ between sides at baseline or change significantly in either group (all p > 0.19). Thus, the condition-specific improvement in flexion acceleration scaling primarily reflected greater pulse-height modulation, that is, improved scaling of initial acceleration amplitude, without a statistically reliable condition-specific change in pulse width or total movement time.

#### Training did not affect acceleration scaling for extension movements

For extension movements of the more-affected arm, peak acceleration also scaled significantly with target amplitude at baseline (β = 20.7 ± 2.3 s⁻², p < 0.001). Scaling did not differ significantly between the Speed (β = 18.0 ± 3.4 s⁻²) and Accuracy (β = 24.0 ± 3.2 s⁻²) groups (p = 0.2), indicating comparable scaling before training. In the Speed group, scaling did not differ between pretraining and the 3-day assessment (Δβ = 6.2 ± 3.8 s⁻², p = 0.22), pretraining and the 1-month assessment (p = 0.52), or the 3-day and 1-month assessments (p = 0.81). Similarly, no significant changes were observed in the Accuracy group (all p ≥ 0.32). No significant group × session interactions were observed for any interval (all p ≥ 0.08).

Exploratory timing analyses likewise provided no consistent evidence of training-related pulse-width modulation for extension. Time to peak velocity did not differ significantly between sides at baseline or change significantly within either group. At baseline, total movement time was slightly shorter on the less-affected side (0.32 ± 0.02 s) than on the more-affected side (0.37 ± 0.025 s; Δ = −0.051 ± 0.024 s, *p* = 0.041), but it did not change significantly across sessions in either group, and no group by session interactions were detected (all p > 0.1).

## Discussion

We hypothesized that, in individuals with chronic stroke, feedforward control in elbow movements on the more-affected side was impaired and that speed-based whole arm training could improve such control. Our results were consistent with both hypotheses. At baseline, peak acceleration scaled with target amplitude on both sides, but scaling was reduced in the more-affected side for both elbow flexion and extension. Speed training increased flexion scaling in 14 of 17 participants in the Speed group. This improvement in scaling at 3 days post-training was large, with the slope more than doubled from baseline to 3 days post-training. Whereas the slope was reduced by 63% on average compared to the less-affected side at baseline, it was only reduced by 23% following training (see Figure 3). In contrast, as hypothesized, Accuracy training did not improve scaling. Because both groups completed the same 2,080 movements, these gains cannot be attributed to movement quantity alone. This result is particularly notable because the Speed group spent 36% less time training and covered 18% less total hand-path distance. Overall, these findings are consistent with a temporary improvement in flexion feedforward control after the speed-emphasized training condition.

Note that our data do not support a complete lack of feedforward control in the chronic phase post-stroke, but rather a reduction in such control. At baseline, initial peak acceleration increased with target amplitude on both sides, indicating that participants retained some capacity to scale initial motor commands to movement extent. However, shallower slopes on the more-affected side indicate that increases in movement distance produced smaller increases in initial peak acceleration. Thus, the primary deficit was not an inability to generate an acceleration pulse, but a reduced ability to grade its magnitude according to planned movement amplitude.

The kinematic timing results support a feedforward interpretation of the initial acceleration peak. Peak acceleration occurred within 80 ms of movement onset across training conditions and sides, consistent with an early feedforward motor command. Similarly, pulse width and total movement time did not change with training. The increase in acceleration scaling therefore did not result from delaying the peak acceleration to use visual feedback corrections (although rapid proprioceptive contributions cannot be ruled out).

At baseline, reduced pulse-height scaling during flexion coincided with a longer time to peak velocity on the more-affected side. This pattern suggests that participants may have prolonged pulse duration to compensate partly for lower acceleration amplitude. Time to peak velocity showed a nonsignificant trend toward shortening at 3 days in the Speed group, returning toward baseline by 1 month, with no group-by-session interaction. Total movement time also remained unchanged across sessions. Consequently, the gain in flexion scaling was driven primarily by acceleration pulse-height modulation rather than reliable changes in pulse duration or overall movement time.

The improved feedforward control following speed training for flexion movements is particularly notable because it transferred from complex whole-arm movement practice to a constrained single-joint task that was not directly practiced during training. Our results are consistent with previous studies indicating that the capacity to update predictive motor commands is partly preserved after chronic stroke. Individuals with chronic stroke adapted to robot-generated force fields and exhibited aftereffects, particularly when forces amplified rather than reduced movement errors (Patton et al. 2006). Adaptation was also observed in response to unpredictable mechanical perturbations, although the influence of prior perturbations and errors was reduced after stroke (Scheidt and Stoeckmann 2007), while split-belt treadmill adaptation produced aftereffects that temporarily reduced gait asymmetry (Reisman et al. 2007). Although findings on motor adaptation after stroke have been inconsistent, a recent meta-analysis showed that motor adaptation is impaired after stroke compared with neurotypical individuals (Abram et al. 2025). Together, these studies indicate that short-term adaptation to experimentally imposed perturbations remains possible after stroke. Importantly, the presence of aftereffects shows that the feedforward command itself was modified by the short-term adaptation processes.

The present findings extend this literature by showing that speed-emphasized practice improved the scaling of peak acceleration occurring within the first 100 ms and transferred to an untrained single-joint task. This provides relatively direct kinematic evidence consistent with modification of the initial feedforward command in chronic stroke, although the effect diminished by 1 month.

This finding may help explain the broader improvements in whole arm reaching movements following the speed training conditions observed in the parent FAST trial (Darmon et al. 2025), as well as in two non-controlled studies (Park et al. 2016; Kantak et al. 2017) in which fast training improved movement time and smoothness, generalized to untrained targets, and persisted for 1 month. Speed training improved movement time, smoothness, velocity-profile symmetry, and, in the FAST trial, the speed–accuracy trade-off during multi-joint reaching. However, these measures combine feedforward planning, feedback correction, and compensatory strategies. These endpoint improvements could potentially arise through several mechanisms, including changes in intersegmental coordination, trunk compensation, feedback corrections, or task-specific strategies. In the present transfer test, movement was mechanically constrained to elbow rotation, visual information about the moving arm was removed at movement onset, and the primary outcome was the initial acceleration scaling. The observed transfer is therefore more consistent with a change in elbow feedforward motor control than with strategies specific to the trained tracks.

Feedback-error learning provides a possible mechanism for this transfer. Feedback controllers generate corrective motor commands in response to movement errors, providing teaching signals to recalibrate feedforward inverse models (Kawato 1999). Rapid multi-joint movements accentuate errors in predicted dynamics, including the timing and magnitude of interaction torques, and may thereby strengthen signals for updating the feedforward control networks. Slower movements minimize dynamics-dependent movement errors and permit online corrections, placing lower demands on feedforward recalibration. Speed practice may improve acceleration scaling by repeatedly exposing the motor system to dynamic errors that feedback corrections alone cannot reduce.

This change may depend on a distributed network involving cortical and cerebellar regions. Lesion studies have demonstrated dissociable neural substrates for online error correction and adaptation after stroke, with left parietal damage specifically impairing the ability to adapt subsequent movements from prior errors (Mutha et al. 2011). Cortico-cerebellar pathways may also contribute to this recalibration process. Although individuals with direct cerebellar lesions were excluded, supratentorial stroke can disrupt descending cortico-ponto-cerebellar fibers, ascending dentato-thalamo-cortical projections, or cerebellar function through crossed cerebellar diaschisis (Infeld et al. 1995; Schulz et al. 2017; Guder et al. 2020). Importantly, the integrity of these cerebellar pathways has also been associated with motor recovery after stroke (Soulard et al. 2020). The consistent response in the Speed group suggests that the capacity for error-driven feedforward updating remained at least partly preserved after stroke. Future work combining behavioral measures with lesion location and cortico-cerebellar structural integrity could determine which neural substrates predict individual gains in acceleration scaling.

Baseline impairment differences may partly explain why Speed training significantly improved flexion but not extension control. Estimated changes in extension in the Speed group were positive but smaller and more variable. Compared with the less-affected side, the baseline scaling deficit was numerically greater for flexion than extension, although this difference was not statistically significant. The greater baseline flexion deficit may therefore have provided more opportunity for improvement, whereas extension scaling was relatively better preserved before training. The relative preservation of extension scaling at baseline may itself reflect prior motor experience. Task-specific upper-extremity movements, particularly outward reaching towards targets, are commonly emphasized during post-stroke rehabilitation (Lang et al. 2009; Winstein et al. 2016; Rozevink et al. 2023; Levin et al. 2023). Moreover, impaired elbow extension and its compensation by increased trunk movement during reaching are well-established features of post-stroke motor control and may have been specifically targeted during rehabilitation (Levin et al. 2016; Roby-Brami et al. 2003; DeJong et al. 2012). Repeated practice aimed at restoring elbow extension while limiting trunk compensation may therefore have contributed to better preservation of extension scaling, leaving less room for improvement with training. However, participants’ prior rehabilitation exposure was not quantified, and this possibility remains speculative.

Lower extension strength may also have constrained scaling gains, as maximal torque was lower for extension than flexion. Reduced force-generating capacity could limit the extent to which participants could increase initial acceleration and its modulation across movement amplitudes, potentially imposing a ceiling on improvements in extension scaling. Abnormal post-stroke flexor synergies could further constrain elbow extension, particularly when shoulder abduction demands are increased (Dewald and Beer 2001; Sukal et al. 2007). However, this effect was likely minimized in the present experimental setup by supporting the arm horizontally and restraining shoulder movement. Furthermore, flexor synergy expression would be expected to impair extension, whereas the baseline scaling deficit was numerically greater in flexion.

Several limitations warrant consideration. First, participants had mild-to-moderate impairment without direct cerebellar damage, limiting generalization to severe paresis or acute stroke. Future studies should determine whether the capacity to modify feedforward scaling is preserved across a broader range of stroke severity and recovery stages. Second, the constrained single-joint task does not capture the full complexity of feedforward control during natural multi-joint reaching, in which the nervous system must anticipate interaction torques generated by motion at adjacent joints (Gribble and Ostry 1999). Although compensatory muscle activity related to interaction torques can still be observed under such conditions (Maeda et al. 2018), future work should determine whether improvements in feedforward scaling at one joint are accompanied by changes in this compensatory muscle activity at neighboring joints. In addition, starting elbow angle covaried with target amplitude, so the scaling slope may include angle-dependent biomechanical effects as well as amplitude-dependent command scaling. Because the same geometry was used at each assessment, this cannot readily explain the within-participant training effect, but future studies should vary amplitude from a common starting angle. Third, because flexion and extension were modeled separately, the absence of an extension effect does not establish direction specificity without a formal direction × group × session test. Finally, gains in flexion scaling were only partly retained, decreasing significantly between 3-day and 1-month post-training. Four training sessions over 1 week were sufficient to induce short-term recalibration but may have been insufficient to stabilize these gains. Longer or distributed training and booster sessions should be investigated as strategies to improve retention.

In conclusion, these results support the view that intensive, speed-emphasized practice of whole-arm movement can improve a fundamental component of joint-level control in an untrained task that minimized inter-joint compensation. Because the task minimized inter-joint compensation, these results support the view that intensive practice can improve a fundamental component of joint-level control in chronic stroke and that gains need not be limited to task-specific compensatory strategies. Incorporating speed demands within safe accuracy boundaries may therefore help target feedforward deficits after stroke, although the partial loss of the effect by 1 month suggests that longer or distributed practice may be needed to stabilize these gains.

## Data Availability

All data produced in the present study are available upon reasonable request to the authors

## Acknowledgments

We thank Hannah Cone and all other clinical staff at Casa Colina Hospital for help with recruitment and data collection, and Alec Roig for technical assistance.

## Funding

This work was funded by grant NIH R21NS120274 to NS.

## Notes

### Competing Interest Statement

The authors have declared no competing interest.

### Clinical Trial

NCT05013762

### Author Declarations

All participants provided written informed consent under a protocol approved by the Institutional Review Board of Casa Colina Hospital (IRB00002372).

## References

Abram, Sabrina J., Jonathan S. Tsay, Heran Yosef, Darcy S. Reisman, and Hyosub E. Kim. 2025. “The Detrimental Effect of Stroke on Motor Adaptation.” Neurorehabilitation and Neural Repair 39 (3): 213–25. 10.1177/15459683241309588.

Beer, R. F., J. P. Dewald, and W. Z. Rymer. 2000. “Deficits in the Coordination of Multijoint Arm Movements in Patients with Hemiparesis: Evidence for Disturbed Control of Limb Dynamics.” Experimental Brain Research 131 (3): 305–19.

Corcos, D. M., G. L. Gottlieb, and G. C. Agarwal. 1989. “Organizing Principles for Single-Joint Movements. II. A Speed-Sensitive Strategy.” Journal of Neurophysiology 62 (2): 358–68. 10.1152/jn.1989.62.2.358.

Darmon, Yannick, Shailesh Kantak, Hannah Cone, et al. 2025. “Speed-Biased Training Temporarily Improves Motor Performance of the Paretic Arm Compared to Accuracy-Biased Training in Chronic Stroke Survivors: The Phase 1 FAST Randomized Clinical Trial.” Neurorehabilitation and Neural Repair 39 (7): 542–54. 10.1177/15459683251331582.

DeJong, Stacey L., Sydney Y. Schaefer, and Catherine E. Lang. 2012. “Need for Speed: Better Movement Quality During Faster Task Performance After Stroke.” Neurorehabilitation and Neural Repair 26 (4): 362–73. 10.1177/1545968311425926.

Dewald, Julius P. A., and Randall F. Beer. 2001. “Abnormal Joint Torque Patterns in the Paretic Upper Limb of Subjects with Hemiparesis.” Muscle & Nerve 24 (2): 273–83. 10.1002/1097-4598(200102)24:2%3C273::AID-MUS130%3E3.0.CO;2-Z.

Dokkum, Liesjet van, Isabelle Hauret, Denis Mottet, Jerome Froger, Julien Métrot, and Isabelle Laffont. 2014. “The Contribution of Kinematics in the Assessment of Upper Limb Motor Recovery Early After Stroke.” Neurorehabilitation and Neural Repair 28 (1): 4–12. 10.1177/1545968313498514.

Gordon, J., and C. Ghez. 1987. “Trajectory Control in Targeted Force Impulses. II. Pulse Height Control.” Experimental Brain Research 67 (2): 241–52. 10.1007/BF00248546.

Gottlieb, G. L., D. M. Corcos, and G. C. Agarwal. 1989. “Organizing Principles for Single-Joint Movements. I. A Speed-Insensitive Strategy.” Journal of Neurophysiology 62 (2): 342–57. 10.1152/jn.1989.62.2.342.

Gottlieb, G. L., D. M. Corcos, G. C. Agarwal, and M. L. Latash. 1990. “Organizing Principles for Single Joint Movements. III. Speed-Insensitive Strategy as a Default.” Journal of Neurophysiology 63 (3): 625–36. 10.1152/jn.1990.63.3.625.

Gribble, Paul L., and David J. Ostry. 1999. “Compensation for Interaction Torques During Single- and Multijoint Limb Movement.” Journal of Neurophysiology 82 (5): 2310–26. 10.1152/jn.1999.82.5.2310.

Guder, Stephanie, Benedikt M. Frey, Winifried Backhaus, et al. 2020. “The Influence of Cortico-Cerebellar Structural Connectivity on Cortical Excitability in Chronic Stroke.” Cerebral Cortex 30 (3): 1330–44. 10.1093/cercor/bhz169.

Infeld, Bernard, Stephen M. Davis, Meir Lichtenstein, Peter J. Mitchell, and John L. Hopper. 1995. “Crossed Cerebellar Diaschisis and Brain Recovery After Stroke.” Stroke 26 (1): 90–95. 10.1161/01.STR.26.1.90.

Johnson, Russell T., Yi Yu, Yannick Darmon, et al. 2026. “Estimating Muscle Parameters via Hierarchical Bayesian Neuromechanics.” bioRxiv, July 9, 2026.07.08.737322. 10.64898/2026.07.08.737322.

Jones, Theresa A. 2017. “Motor Compensation and Its Effects on Neural Reorganization after Stroke.” Nature Reviews Neuroscience 18 (5): 267–80. 10.1038/nrn.2017.26.

Kamper, Derek G., Alicia N. McKenna-Cole, Leonard E. Kahn, and David J. Reinkensmeyer. 2002. “Alterations in Reaching after Stroke and Their Relation to Movement Direction and Impairment Severity.” Archives of Physical Medicine and Rehabilitation 83 (5): 702–7. 10.1053/apmr.2002.32446.

Kantak, Shailesh S., Nazaneen Zahedi, and Robert McGrath. 2017. “Complex Skill Training Transfers to Improved Performance and Control of Simpler Tasks After Stroke.” Physical Therapy 97 (7): 718–28. 10.1093/ptj/pzx042.

Kawato, Mitsuo. 1999. “Internal Models for Motor Control and Trajectory Planning.” Current Opinion in Neurobiology 9 (6): 718–27. 10.1016/S0959-4388(99)00028-8.

Kawato, Mitsuo, and Hiroaki Gomi. 1992. “A Computational Model of Four Regions of the Cerebellum Based on Feedback-Error Learning.” Biological Cybernetics 68 (2): 95–103. 10.1007/BF00201431.

Laczko, Jozsef, Robert A. Scheidt, Lucia S. Simo, and Davide Piovesan. 2017. “Inter-Joint Coordination Deficits Revealed in the Decomposition of Endpoint Jerk During Goal-Directed Arm Movement After Stroke.” IEEE Transactions on Neural Systems and Rehabilitation Engineering 25 (7): 798–810. 10.1109/TNSRE.2017.2652393.

Lang, Catherine E., Jillian R. MacDonald, Darcy S. Reisman, et al. 2009. “Observation of Amounts of Movement Practice Provided During Stroke Rehabilitation.” Archives of Physical Medicine and Rehabilitation 90 (10): 1692–98. 10.1016/j.apmr.2009.04.005.

Levin, Mindy F., Sigal Berman, Neta Weiss, et al. 2023. “ENHANCE Proof-of-Concept Three-Arm Randomized Trial: Effects of Reaching Training of the Hemiparetic Upper Limb Restricted to the Spasticity-Free Elbow Range.” Scientific Reports 13 (1): 22934. 10.1038/s41598-023-49974-6.

Levin, Mindy F., Jeffrey A. Kleim, and Steven L. Wolf. 2009. “What Do Motor ‘Recovery’ and ‘Compensation’ Mean in Patients Following Stroke?” Neurorehabilitation and Neural Repair 23 (4): 313–19. 10.1177/1545968308328727.

Levin, Mindy F., Dario G. Liebermann, Yisrael Parmet, and Sigal Berman. 2016. “Compensatory Versus Noncompensatory Shoulder Movements Used for Reaching in Stroke.” Neurorehabilitation and Neural Repair 30 (7): 635–46. 10.1177/1545968315613863.

Lum, Peter S., Sara Mulroy, Richard L. Amdur, Philip Requejo, Boris I. Prilutsky, and Alexander W. Dromerick. 2009. “Gains in Upper Extremity Function After Stroke via Recovery or Compensation: Potential Differential Effects on Amount of Real-World Limb Use.” Topics in Stroke Rehabilitation 16 (4): 237–53. 10.1310/tsr1604-237.

Maeda, Rodrigo S., Tyler Cluff, Paul L. Gribble, and J. Andrew Pruszynski. 2018. “Feedforward and Feedback Control Share an Internal Model of the Arm’s Dynamics.” Research Articles. Journal of Neuroscience 38 (49): 10505–14. 10.1523/JNEUROSCI.1709-18.2018.

McCrea, Patrick H., and Janice J. Eng. 2005. “Consequences of Increased Neuromotor Noise for Reaching Movements in Persons with Stroke.” Experimental Brain Research 162 (1): 70–77. 10.1007/s00221-004-2106-8.

Mottet, Denis, Liesjet Elisabeth Henriette van Dokkum, Jérôme Froger, Abdelkader Gouaïch, and Isabelle Laffont. 2017. “Trajectory Formation Principles Are the Same after Mild or Moderate Stroke.” PLOS ONE 12 (3): e0173674. 10.1371/journal.pone.0173674.

Murphy, Margit Alt, Carin Willén, and Katharina S. Sunnerhagen. 2011. “Kinematic Variables Quantifying Upper-Extremity Performance After Stroke During Reaching and Drinking From a Glass.” Neurorehabilitation and Neural Repair 25 (1): 71–80. 10.1177/1545968310370748.

Mutha, Pratik K., Robert L. Sainburg, and Kathleen Y. Haaland. 2011a. “Left Parietal Regions Are Critical for Adaptive Visuomotor Control.” The Journal of Neuroscience: The Official Journal of the Society for Neuroscience 31 (19): 6972–81. 10.1523/JNEUROSCI.6432-10.2011.

Nibras, Nadir, Chang Liu, Denis Mottet, et al. 2021. “Dissociating Sensorimotor Recovery and Compensation During Exoskeleton Training Following Stroke.” Frontiers in Human Neuroscience 15 (April): 645021. 10.3389/fnhum.2021.645021.

Park, Hyeshin, Sujin Kim, Carolee J. Winstein, James Gordon, and Nicolas Schweighofer. 2016. “Short-Duration and Intensive Training Improves Long-Term Reaching Performance in Individuals With Chronic Stroke.” Neurorehabilitation and Neural Repair 30 (6): 551–61. 10.1177/1545968315606990.

Patton, James L., Mary Ellen Stoykov, Mark Kovic, and Ferdinando A. Mussa-Ivaldi. 2006. “Evaluation of Robotic Training Forces That Either Enhance or Reduce Error in Chronic Hemiparetic Stroke Survivors.” Experimental Brain Research 168 (3): 368–83. 10.1007/s00221-005-0097-8.

Raj, Sandesh, Natalia Dounskaia, William W. Clark, and Amit Sethi. 2020. “Effect of Stroke on Joint Control during Reach-to-Grasp: A Preliminary Study.” Journal of Motor Behavior 52 (3): 294–310. 10.1080/00222895.2019.1615861.

Reinkensmeyer, David J., Mario G. Iobbi, Leonard E. Kahn, Derek G. Kamper, and Craig D. Takahashi. 2003. “Modeling Reaching Impairment After Stroke Using a Population Vector Model of Movement Control That Incorporates Neural Firing-Rate Variability.” Neural Computation 15 (11): 2619–42. 10.1162/089976603322385090.

Reisman, Darcy S., Robert Wityk, Kenneth Silver, and Amy J. Bastian. 2007. “Locomotor Adaptation on a Split-Belt Treadmill Can Improve Walking Symmetry Post-Stroke.” Brain 130 (7): 1861–72. 10.1093/brain/awm035.

Roby-Brami, Agnès, Stéphane Jacobs, Nezha Bennis, and Mindy F. Levin. 2003. “Hand Orientation for Grasping and Arm Joint Rotation Patterns in Healthy Subjects and Hemiparetic Stroke Patients.” Brain Research 969 (1): 217–29. 10.1016/S0006-8993(03)02334-5.

Rozevink, Samantha G., Juha M. Hijmans, Koen A. Horstink, and Corry K. van der Sluis. 2023. “Effectiveness of Task-Specific Training Using Assistive Devices and Task-Specific Usual Care on Upper Limb Performance after Stroke: A Systematic Review and Meta-Analysis.” Disability and Rehabilitation. Assistive Technology 18 (7): 1245–58. 10.1080/17483107.2021.2001061.

Sainburg, Robert L., and Sydney Y. Schaefer. 2004. “Interlimb Differences in Control of Movement Extent.” Journal of Neurophysiology 92 (3): 1374–83. 10.1152/jn.00181.2004.

Scheidt, Robert A., and Tina Stoeckmann. 2007. “Reach Adaptation and Final Position Control amid Environmental Uncertainty after Stroke.” Journal of Neurophysiology 97 (4): 2824–36. 10.1152/jn.00870.2006.

Schulz, Robert, Benedikt M. Frey, Philipp Koch, et al. 2017. “Cortico-Cerebellar Structural Connectivity Is Related to Residual Motor Output in Chronic Stroke.” Cerebral Cortex 27 (1): 635–45. 10.1093/cercor/bhv251.

Schweighofer, Nicolas, Michael A. Arbib, and Mitsuo Kawato. 1998. “Role of the Cerebellum in Reaching Movements in Humans. I. Distributed Inverse Dynamics Control.” European Journal of Neuroscience 10 (1): 86–94. 10.1046/j.1460-9568.1998.00006.x.

Schweighofer, Nicolas, Jacob Spoelstra, Michael A. Arbib, and Mitsuo Kawato. 1998. “Role of the Cerebellum in Reaching Movements in Humans. II. A Neural Model of the Intermediate Cerebellum.” European Journal of Neuroscience 10 (1): 95–105. 10.1046/j.1460-9568.1998.00007.x.

Soulard, Julie, Coline Huber, Sebastien Baillieul, et al. 2020. “Motor Tract Integrity Predicts Walking Recovery: A Diffusion MRI Study in Subacute Stroke.” Article. Neurology 94 (6): e583–93. 10.1212/WNL.0000000000008755.

Stewart, Jill Campbell, James Gordon, and Carolee J. Winstein. 2014. “Control of Reach Extent with the Paretic and Nonparetic Arms after Unilateral Sensorimotor Stroke: Kinematic Differences Based on Side of Brain Damage.” Experimental Brain Research 232 (7): 2407–19. 10.1007/s00221-014-3938-5.

Sukal, Theresa M., Michael D. Ellis, and Julius P. A. Dewald. 2007. “Shoulder Abduction-Induced Reductions in Reaching Work Area Following Hemiparetic Stroke: Neuroscientific Implications.” Experimental Brain Research 183 (2): 215–23. 10.1007/s00221-007-1029-6.

Winstein, Carolee J., Steven L. Wolf, Alexander W. Dromerick, et al. 2016. “Effect of a Task-Oriented Rehabilitation Program on Upper Extremity Recovery Following Motor Stroke: The ICARE Randomized Clinical Trial.” JAMA 315 (6): 571–81. 10.1001/jama.2016.0276.

